# Environmental surveillance for SARS-CoV-2 for outbreak detection in hospital: A single centre prospective study

**DOI:** 10.1101/2023.08.28.23294549

**Authors:** Prachi Ray, Bryant Lim, Katarina Zorcic, Jennie Johnstone, Aaron Hinz, Alexandra M.A. Hicks, Alex Wong, Derek R. MacFadden, Caroline Nott, Lucas Castellani, Rees Kassen, Michael Fralick

## Abstract

Identifying COVID-19 outbreaks in hospitals at an early stage requires active surveillance. Our objective was to assess whether floor swabs correlated with COVID-19 outbreak status in hospital. We swabbed the floors of an inpatient ward at Mount Sinai Hospital for 32 weeks, from October 31, 2022 to June 15, 2023 and RT-qPCR analysis provided a quantification cycle of detection for each positive swab. 182 swabs were processed for SARS CoV-2, of which 98.4% were positive. Two COVID-19 outbreaks were declared during the study period. The median viral copy number was 210 (IQR, 49 to 1018) during non-outbreak periods and 653 (IQR, 300 to 1754) during outbreak periods. Analyzing the number of viral copies of SARS-CoV-2, instead of percentage positivity, gave a clearer view of changes in outbreak status over time, thereby illustrating the benefits of this approach to monitor pathogen load in hospital settings.

## BACKGROUND

Early identification of COVID-19 outbreaks in hospitals requires active surveillance. Typically, this involves daily systematic review of symptoms in hospitalized patients. However, this approach does not account for pre-symptomatic, asymptomatic, or unrecognized symptoms in admitted patients, and COVID-19 outbreaks continue to pose a challenge in healthcare settings. Prior work suggests that environmental surveillance (e.g., wastewater surveillance, built environment sampling) may be an effective method for active surveillance.^1–4^ Our research team previously conducted a 14-month study in which we swabbed the floors once weekly at 10 long-term care homes and identified that the percentage of floor swabs positive for SARS-CoV-2 was predictive of an impending COVID-19 outbreak (AUC 0.84).^1^ During non-outbreak periods, 22% of floor swabs were positive for SARS-CoV-2; during COVID-19 outbreaks, floor swab positivity rose to 54%. Floor swab positivity also mirrored outbreak activity: as outbreaks worsened, the percentage of positive floor swabs rose, and as outbreaks resolved, the percentage of positive floor swabs fell. This study’s objective was to assess if floor swab positivity follows a similar pattern before and after a COVID-19 outbreak in the inpatient unit of a hospital.

## METHODS

We conducted a 32-week prospective study at Mount Sinai Hospital, a tertiary care hospital located in Toronto, Canada. Beginning in Fall 2022, we swabbed the hallways of one of the general internal medicine floors (see Appendix for floor map). This floor has 16 patient rooms. Of these, 6 are single rooms, 9 are double rooms with 2-patient capacity, and 1 is a 4-patient room. The maximum patient capacity on the ward is 28 patients. Swabs were taken in the 3 main hallways, which are each 2.5 metres in width and of varying lengths (37 metres, 27 metres, and 9 metres). There is no dedicated COVID-19 unit on this floor, but patients with COVID-19 can be admitted to this ward. COVID-19 mitigation measures in place during the study period included universal masking policies, PCR testing of symptomatic staff, and PCR testing of all patients upon their admission to the hospital.

Comprehensive weekly sampling of the high traffic areas along all hallways outside patient rooms and staff working areas was conducted, comprising approximately 10 swabs per week, at approximately 5-metre intervals (see Appendix). Floor swabs were performed using previously validated protocols^5^ and involved swabbing across 5 × 5 centimetre areas using the P-208 Environmental Surface Collection Prototype kit from DNA Genotek. The quantitative reverse transcriptase-PCR (RT-qPCR) results provided a quantification cycle (Cq) value for each positive swab, which was subsequently converted into copy numbers of the virus using a standard curve detailed in previously validated methods.^5^

We collected floor swab samples from October 31, 2022 to June 15, 2023. This start date was selected because there was a COVID-19 outbreak in the chosen inpatient unit that ended on October 28, 2022.

An outbreak was defined by Sinai Health as two cases of COVID-19 within a 14-day period where both could reasonably have been acquired in the hospital. A suspected outbreak was defined as two nosocomial cases of COVID-19 with no link to one another, identified within 7 days. A suspected outbreak only leads to a declared outbreak if additional COVID-19 cases are evident after 5 to 7 days of calling the suspected outbreak. During the study period, the following two COVID-19 outbreaks were declared on the ward: December 20, 2022 to January 8, 2023 (19 days) and March 17, 2023 to March 30, 2023 (13 days). A suspected outbreak was declared on April 11, 2023 but did not lead to a declared outbreak.

Descriptive statistics for the percentage of positive floor swabs and copy numbers of the virus over time were reported. Because the copy numbers were not normally distributed, non-parametric tests were performed to compare copy numbers before and after an outbreak.

## RESULTS

During the 32-week time period, 182 swabs were collected on 21 separate days. The overall floor swab positivity for SARS-CoV-2 was 98.4%. During COVID-19 outbreak periods, the floor swab positivity was 100%, and during non-outbreak periods it was 98.0%. In our previous work, percentage positivity was used as the primary predictor of outbreak status.^1,2,5^ For this study, because positivity was near 100%, we also assessed viral copy number as a predictor. The median viral copy number throughout the duration of the study was 279 (interquartile range [IQR], 68 to 1230). During COVID-19 outbreak periods, the median viral copy number was 653 (IQR, 300 to 1754) and during non-outbreak periods it was 210 (IQR, 49 to 1018). In Figure 1, we provide a graphical representation of how viral copy numbers changed over time. Two COVID-19 outbreaks occurred during the 32-week period, and an increase in viral material (i.e., number of copies) is evident prior to the declaration of each outbreak, and number of copies remains high during the outbreak period (Figure 1).

**Figure 1.**
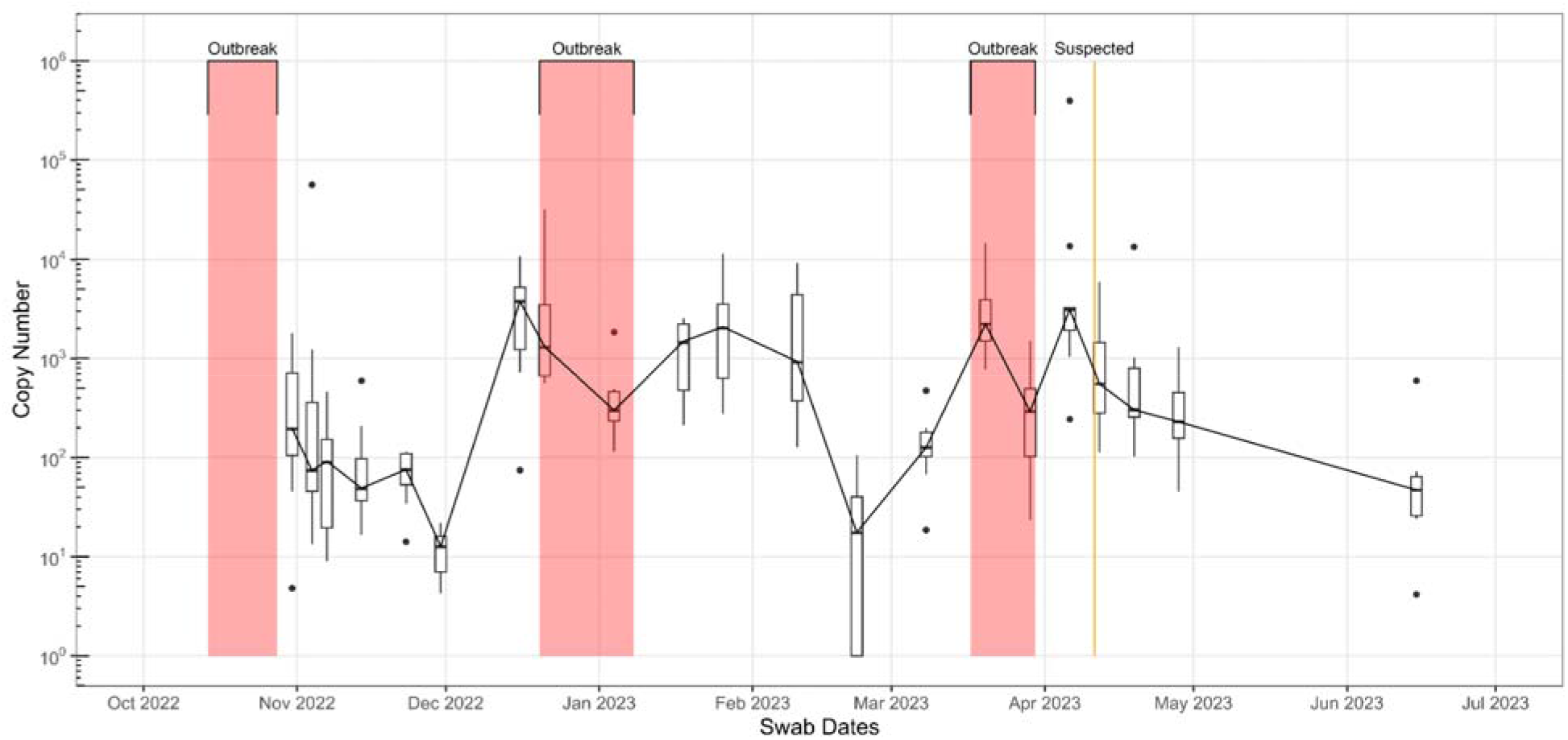
SARS-CoV-2 viral copy numbers on the floors of the inpatient ward on log scale. One outbreak ended just prior to the study period. Two outbreaks and one suspected outbreak occurred during the 32-week period, highlighted in red and orange respectively.

## DISCUSSION

Previously, we and others have proposed the use of floor swab positivity to predict COVID-19 outbreaks. However, this approach is unsuitable in settings with persistently high swab positivities (i.e., approaching 100%). Our prospective study demonstrates that while the percentage floor swabs positive for SARS-CoV-2 was near 100% regardless of the presence of a COVID-19 outbreak, the number of detected viral copies mirrored the COVID-19 outbreak status. In particular, we identified that viral copy numbers were typically highest at the start of an outbreak and fell toward the end of an outbreak.

Our study aligns with previous viral detection studies conducted in patient care settings.^6,7^ One of the earliest environmental swabbing studies conducted in Wuhan, China in 2020 found a high rate of positivity (70%) of SARS-CoV-2 from floor swabs taken in an intensive care unit.^8^ Studies conducted in medical centres and tertiary care hospitals have found that the likelihood of detecting SARS-CoV-2 on swabs depends on surface type; specifically, floors were shown to have the highest probability of SARS-CoV-2 detection, compared to other surfaces such as walls, patient bed rails, or doorknobs.^5,9^ Our results support the notion that floor swabs can be used for the detection of SARS-CoV-2, and also illustrate that the viral load in the built environment vary depending on the outbreak status in patient care settings.

Because nearly 100% of our floor swabs were positive for SARS-CoV-2 regardless of outbreak status on the ward, percentage positivity would be an ineffective method of detecting changes in SARS-CoV-2 burden in the hospital setting. This differs from our previous study conducted in long-term care homes in which floor swab positivity was significantly different between outbreak and non-outbreak periods. Unlike hospital settings, however, long-term care homes strive to minimize the number of COVID-19–positive residents—typically, if a resident is infected, they are transferred to hospital. Patient wards, on the other hand, regularly admit patients with COVID-19, which likely explains the consistent detection of SARS-CoV-2 on the hospital floor. By analyzing the number of viral copies of SARS-CoV-2 instead of percentage positivity of the floor swabs, we obtained a clearer view of changes in viral load over time.

An important limitation of our study is that it was a single-centre evaluation. However, the results align with our prior multicentre prospective study that included 23 outbreaks over a 14-month period across 10 long-term care homes.^1^ To increase generalizability, it will be important to identify whether our observed trends also apply to patient care settings with fewer COVID-19 mitigation measures, and/or during periods of increased burden of other respiratory viruses. Future studies will be needed to validate our findings and assess whether environmental surveillance can help inform infection control measures in hospitals.

## Data Availability

All data produced in the present study are available upon reasonable request to the authors.

**Appendix Figure 1.**
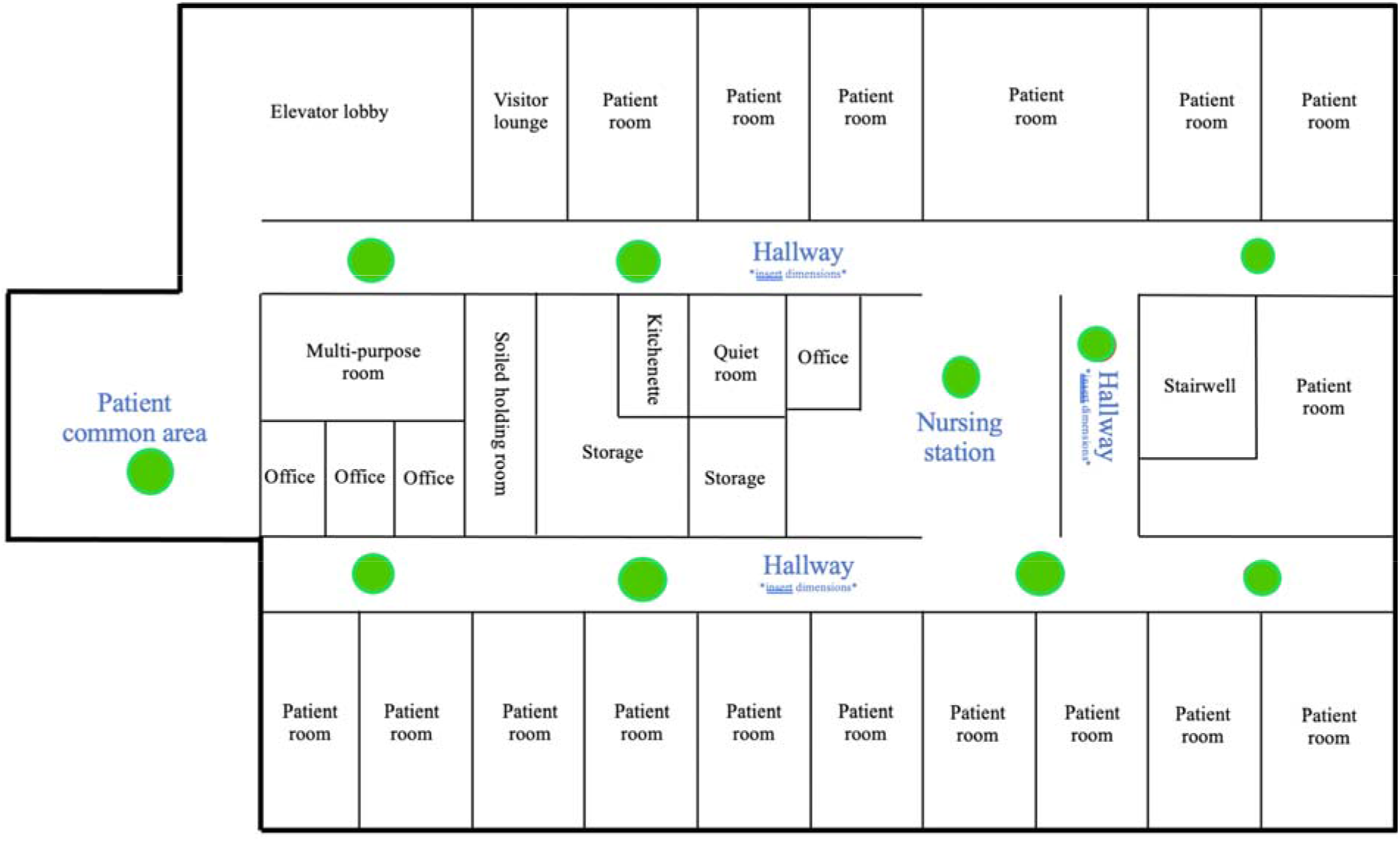
Floor map for the internal medicine ward where swabbing occurred. Green circles indicate swabbing locations (i.e., floors in the hallways, nursing station, and patient common area).

## References

1. Fralick M, Nott C, Moggridge J, et al. Detection of covid-19 outbreaks using built environment testing for SARS-CoV-2. NEJM Evid. 2023;2(3). doi:10.1056/evidoa2200203

2. Fralick M, Burella M, Hinz A, et al. The spatial and temporal distribution of SARS-CoV-2 from the built environment of COVID-19 patient rooms: A multicentre prospective study. PLoS One. 2023;18(3):e0282489.

3. Manuel DG, Delatolla R, Fisman DN, et al. The Role of Wastewater Testing for SARS-CoV-2 Surveillance. Ontario COVID-19 Science Advisory Table; 2021. doi:10.47326/ocsat.2021.02.40.1.0

4. Coil DA, Albertson T, Banerjee S, et al. SARS-CoV-2 detection and genomic sequencing from hospital surface samples collected at UC Davis. PLoS One. 2021;16(6):e0253578.

5. Hinz A, Xing L, Doukhanine E, et al. SARS-CoV-2 detection from the built environment and wastewater and its use for hospital surveillance. Facets (Ott). 2022;7:82–97.

6. Minich JJ, Ali F, Marotz C, et al. Feasibility of using alternative swabs and storage solutions for paired SARS-CoV-2 detection and microbiome analysis in the hospital environment. Microbiome. 2021;9(1):25.

7. Ong SWX, Tan YK, Chia PY, et al. Air, Surface Environmental, and Personal Protective Equipment Contamination by Severe Acute Respiratory Syndrome Coronavirus 2 (SARS-CoV-2) From a Symptomatic Patient. JAMA. 2020;323(16):1610–1612.

8. Guo ZD, Wang ZY, Zhang SF, et al. Aerosol and Surface Distribution of Severe Acute Respiratory Syndrome Coronavirus 2 in Hospital Wards, Wuhan, China, 2020. Emerg Infect Dis. 2020;26(7):1583–1591.

9. Ziegler MJ, Huang E, Bekele S, et al. Spatial and temporal effects on severe acute respiratory coronavirus virus 2 (SARS-CoV-2) contamination of the healthcare environment. Infect Control Hosp Epidemiol. 2022;43(12):1773–1778.

